# Disentangling Type 2 Diabetes Risk and Comorbidity Using Genomic Structural Equation Modeling

**DOI:** 10.64898/2026.01.26.26344825

**Authors:** Merli Koitmäe, Kristi Läll, Märt Möls, Andrew P. Morris, Krista Fischer, Reedik Mägi

## Abstract

Type 2 diabetes (T2D) is a genetically and clinically heterogeneous disorder influenced by metabolic, lifestyle, and cardiometabolic factors. Understanding the shared and distinct genetic architecture underlying T2D and its upstream risk traits is critical for improved risk prediction and stratified intervention. We applied genomic structural equation modeling (SEM) to genome-wide association study (GWAS) summary statistics for eight T2D-related phenotypes: gestational diabetes, hypertension, glycated hemoglobin, fasting glucose, fasting insulin, insulin sensitivity, body mass index, and waist-to-hip ratio. Exploratory and confirmatory factor analyses identified three latent genetic factors representing glycemic regulation, insulin resistance and cardiometabolic risk, and obesity and lifestyle-related traits. Polygenic scores (PGSs) derived from multivariate GWAS of these latent factors predicted incident T2D in the Estonian Biobank, capturing overlapping yet distinct risk profiles compared with conventional metabolic PGSs. Phenome-wide and comorbidity analyses revealed that each latent factor PGS associated with specific T2D-related complications and provided broader or distinct associations compared with conventional metabolic PGSs, with the obesity and lifestyle-related factor showing the widest impact. These findings illustrate how multivariate genetic approaches can disentangle the biological heterogeneity of T2D, refine polygenic risk prediction, and reveal mechanistic pathways driving divergent patterns of disease and comorbidity.

## Introduction

Diabetes mellitus is a major global health burden. The IDF Diabetes Atlas estimates that 589 million adults are living with diabetes worldwide, with this number projected to increase to 853 million adults by 2050 (1). More than 90% of individuals living with diabetes mellitus are affected by type 2 diabetes (T2D), a condition characterised by hyperglycaemia, impaired insulin secretion, and insulin resistance. Beyond its widespread occurrence, T2D imposes a substantial public health burden due to its chronic nature and wide range of complications, including cardiovascular disease, nephropathy, neuropathy, and retinopathy (2).

T2D results from complex interplay between lifestyle, environmental, and genetic risk factors, with heritability estimated to be 69% for individuals aged between 35 and 60 years old (3). The most recent multi-ancestry genome-wide association study (GWAS) meta-analysis of T2D, which aggregated data from 2,535,601 individuals (428,452 T2D cases), identified 1,289 independent association signals (mapping to 611 loci) at genome-wide significance (*P*<5x10^-8^) (4). These loci point to diverse biological pathways leading to the development of T2D that include pancreatic development, beta-cell function and insulin secretion, glucose and lipid metabolism, WNT signalling and inflammation, which lead to considerable pathophysiological heterogeneity in individuals with T2D leading to variability in disease trajectories including age of onset, development of complications, and response to treatment (5–8). Because many upstream cardiometabolic traits such as obesity, hypertension, impaired insulin action, and altered glucose homeostasis strongly predispose individuals to T2D, clarifying their shared and distinct etiological contributions is essential for improving disease prediction and prevention.

To advance precision medicine in T2D, alternative approaches have been used to define disease subtypes using clinical, biomarker, and genetic data (6,7,9). A landmark data-driven cluster analysis of adult-onset diabetes by Ahlqvist et al. identified reproducible subgroups characterized by severe insulin-deficient diabetes, severe insulin-resistant diabetes, mild obesity-related diabetes, mild age-related diabetes, and autoimmune diabetes, each with specific risks for complications and differing clinical trajectories (9). These findings highlight that the heterogeneity of T2D reflects underlying variation in metabolic dysfunction, insulin secretion, insulin sensitivity, adiposity, and related pathophysiological axes. Studying both overt T2D and upstream quantitative traits such as fasting glucose, fasting insulin, insulin sensitivity, body mass index (BMI), and hypertension can therefore reveal the genetic and physiological structure that gives rise to this heterogeneity.

A promising approach to understanding aetiological heterogeneity is to cluster T2D-associated genetic variants according to their profile of associations with diabetes-related cardiometabolic traits. The expectation is that each cluster of genetic variants represents a pathway-specific process leading to disease, and the combination of cluster-specific polygenic scores (PGSs) can be used to predict disease onset and progression (10). Multiple strategies for clustering have been employed, including “hard clustering” (in which each genetic variant is assigned to exactly one cluster) and “soft clustering” (in which variants can be assigned to multiple clusters, or to no clusters at all). Irrespective of the method, selection of genetic variants, or range of cardiometabolic traits used for the clustering, five pathway-specific clusters have been consistently identified, which reflect beta-cell dysfunction (with positive or negative association with proinsulin), obesity, lipodystrophy, and liver/lipid metabolism (5–8). Pathway-specific partitioned PGSs for the obesity cluster have been reported to be strongly associated with increased risk of a range of macro- and microvascular complications, whilst those for the lipodystrophy cluster have been reported to be associated with increased risk of coronary artery disease. Together, these findings indicate that distinct metabolic and genetic pathways drive divergent comorbid trajectories among T2D subgroups, underscoring the need for stratified approaches to risk prediction and intervention.

Genomic structural equation modeling (genomic SEM) offers a powerful framework for characterizing shared genetic architecture across correlated traits and for identifying latent factors that represent core biological dimensions. By integrating GWAS summary statistics into a multivariate model, genomic SEM enables joint estimation of genetic correlations, latent trait structures, and multivariate GWAS associations that cannot be captured by single-trait analyses (11). Applications of genomic SEM have revealed fundamental axes of genetic liability across psychiatric disorders (12), cardiometabolic traits (13) and metabolic syndrome (14), demonstrating substantial gains in interpretability and locus discovery compared to univariate approaches. Our aim is to apply genomic SEM to T2D-related phenotypes to identify latent cardiometabolic components reflecting glycemic regulation, insulin resistance, adiposity, and metabolic stress, improving both biological insight, downstream polygenic prediction and cluster T2D patients to identify their unique risk for comorbidities.

## Material and Methods

### Data

#### Definition of phenotypes

We focused on eight phenotypes that capture complementary aspects of T2D risk and onset: gestational diabetes (GDM), hypertension (Hyp), glycated hemoglobin (HbA1c), fasting insulin (FI), fasting glucose (FG), insulin sensitivity (IS), BMI and waist to hip ratio (WHR) adjusted for BMI. Some phenotypes (HbA1c and FG) are integral to T2D diagnosis, while others (FI, IS, BMI, WHR) reflect upstream metabolic and adiposity-related risk factors that typically precede disease. GDM and hypertension further highlight systemic vulnerability, as they often emerge before T2D and strongly predict future progression (2).

#### Selection of GWAS

For the eight phenotypes of interest, we selected summary statistics from the largest available GWAS conducted in European-ancestry cohorts. To identify the most suitable summary statistics, the GWAS Catalog (15), PubMed (16), and relevant consortium websites, including MAGIC (17) and DIAGRAM (18), were screened.

#### Estonian biobank data

Estonian biobank (19,20) (EstBB) includes 212,000 mainly European-ancestry participants. All EstBB participants were genotyped at the Core Genotyping Lab of the Institute of Genomics, University of Tartu, using Illumina Global Screening Array v1.0, v2.0, and v3.0. Genotypes were called and PLINK files were generated using Illumina GenomeStudio v2.0.4. Individuals were excluded if their call-rate was < 95%, if they were outliers in heterozygosity (> 3 SD from the mean), or if sex inferred from X chromosome heterozygosity did not match the reported sex (21). Variants were filtered prior to imputation for call-rate < 95%, Hardy–Weinberg equilibrium p-value < 1×10LJLJ (autosomal variants only), and minor allele frequency < 1%. Variant positions were lifted over from GRCh37 to GRCh38 using Picard. Phasing was performed with Beagle v5.4 (22), and imputation was carried out using Beagle v5.4 (beagle.22Jul22.46e.jar) in batches of 5,000 variants. A population-specific reference panel consisting of 2,695 WGS samples (21) was used for imputation along with standard Beagle hg38 recombination maps. Based on principal component analysis, samples of non-European ancestry were removed. Duplicate and monozygous twin detection was performed with KING 2.2.7 (23) and one sample from each duplicate pair was excluded.

EstBB participants’ records are regularly updated by retrieving data from the national health databases of the Estonian Health Insurance Fund and the National Health Information System. This allowed us to complement the available genotype data with clinically relevant phenotypic information. Specifically, we obtained the age at diagnosis of T2D (ICD-10 code E11), as well as the age at diagnosis of common T2D-related comorbidities, including disorders of lipoprotein metabolism and other lipidaemias (E78), mononeuropathies of upper limb (G56), retinal disorders (H36), glaucoma (H40), hypertensive heart disease (I11), hypertensive heart and renal disease (I13), heart failure (I50), other diseases of liver (K76), cholelithiasis (K80), gout (M10), glomerular diseases (N08), and chronic renal failure (N18) (24–29).

#### Data analysis Factor analysis

To compute the genetic correlation matrix and the corresponding sampling covariance matrices of GWAS parameters for the eight phenotypes included in the analysis the multivariable extension of linkage disequilibrium (LD) score regression (11) (LDSC) implemented in the ldsc function in GenomicSEM v0.0.5 (30) was used. The HapMap3 reference panel was used for LD estimation. All GWASs were filtered with imputation quality filter 0.8 (if present) and a minor allele frequency (MAF) filter of 1%.

Exploratory factor analysis (EFA) was conducted using promax rotation. Confirmatory factor analysis (CFA) was then performed using the usermodel function in GenomicSEM v0.0.5. All latent variables were standardized using unit variance identification, and model parameters were estimated using the diagonally weighted least squares (DWLS) method (11).

The genetic factor model was constructed based on the EFA results, using indicators with factor loadings ≥ 0.25. Model fit was assessed using the Comparative Fit Index (CFI) and the Standardized Root Mean Residual (SRMR). The model was considered to have a good fit if CFI > 0.95 and SRMR < 0.08 (31).

#### Multivariate GWAS with Genomic SEM

Associations between SNPs and latent factors derived from confirmatory factor analysis were estimated in multivariate GWAS using the userGWAS function in the GenomicSEM package (v0.0.5). All latent factors were constrained to have unit variance, and model estimation was performed using the DWLS method. (11) GWAS summary statistics used in the multivariate GWAS were filtered to include SNPs with MAF > 1%, and the 1000 Genomes European reference panel was used as a reference. The visualization of the results was performed using Manhattan plots, while annotating the top SNP in each chromosome. The corresponding nearest genes were assigned using dbSNP database (32). *n_eff_* for the latent factor was estimated based on the equation provided by Mallard et al (33) and SNPs with MAF between 10% and 40% were used for this to produce more stable estimates.

#### PGS analysis

PGSs were computed using PRS-continuous shrinkage (PRS-CS) (34) for the three latent factors identified through Genomic SEM, as well as for type 2 diabetes (T2D) and the three primary phenotypes most strongly influenced by each factor. As the GWAS summary statistics for FG and FI included a small subset of Estonian Biobank participants, these participants were excluded from all subsequent analyses to avoid potential sample overlap. For T2D, we used the polygenic score PGS002026, originally developed by Privé et al. (35), and made available through the PGS Catalog (36). We analysed the effect of the PGSs on incident T2D using Cox PH models using age as timescale and restricted the analysis for people over 40. All models were adjusted for sex and 10 genetic principal components (PCs). PGS overlaps were visualized using Venn diagrams.

#### Phenome-wide association study (PheWAS) with PGS

PheWAS analysis was performed using latent factor PGSs, computed using multivariate GWAS results from Genomic SEM, as well as PGSs for FG, FI and BMI. A logistic regression model was fitted to 1,942 disease phenotypes, defined by ICD-10 codes, adjusted for sex, year of birth and ten genetic PCs. All relatives with identity by descent greater than 20% in the Estonian Biobank were excluded for this analysis. The results were visualized using phenotypeManhattan function in the PheWAS package (37).

#### Comorbidity analyses

Comorbidity analyses were conducted using Cox proportional hazards models with age as timescale. The follow-up time began at the age of T2D diagnosis and ended at either the age of the first comorbidity diagnosis or the age at last electronic health record (EHR) linkage for censored individuals. The comorbidities were chosen to be the known comorbidities of T2D as described above. All models were adjusted for sex and the first 10 genetic PCs. The analysis was restricted to individuals, whose age was at least 40 at the diagnosis of T2D as some of the diagnoses in younger age are likely to be misclassified T1D diagnoses. Kaplan–Meier curves were used to visualize the time from T2D diagnosis to the onset of comorbid conditions. For this, we divided the EstBB participants into three categories based on the PGSs: top 20%, middle 60% and bottom 20%.The comorbid conditions listed previously were selected based on PGS-PheWAS results and supporting literature. All analyses were implemented in R using the survival package (38) and visualized using ggplot2 package (39).

## Results

### Selection of GWAS

We collected summary statistics from GWAS for eight established risk factors associated with T2D: GDM, Hyp, HbA1c, FI, FG, IS, BMI, and WHR (adjusted for BMI). The GWAS sample sizes ranged from 53,710 to 806,834. The original GWAS studies and sample sizes of each dataset are provided in Supplementary Table 1.

### Genetic correlations between T2D risk factors

Genetic correlations among the eight T2D risk factors ranged from −0.645 (IS–FI) to 0.537 (GDM–FG) (Figure 1). The correlations were mainly positive, indicative of shared genetic architecture, whereas IS showed negative correlations with the other traits, consistent with its protective role: greater insulin sensitivity is genetically associated with lower insulin levels, improved glycaemic control, and reduced T2D risk.

**Figure 1.**
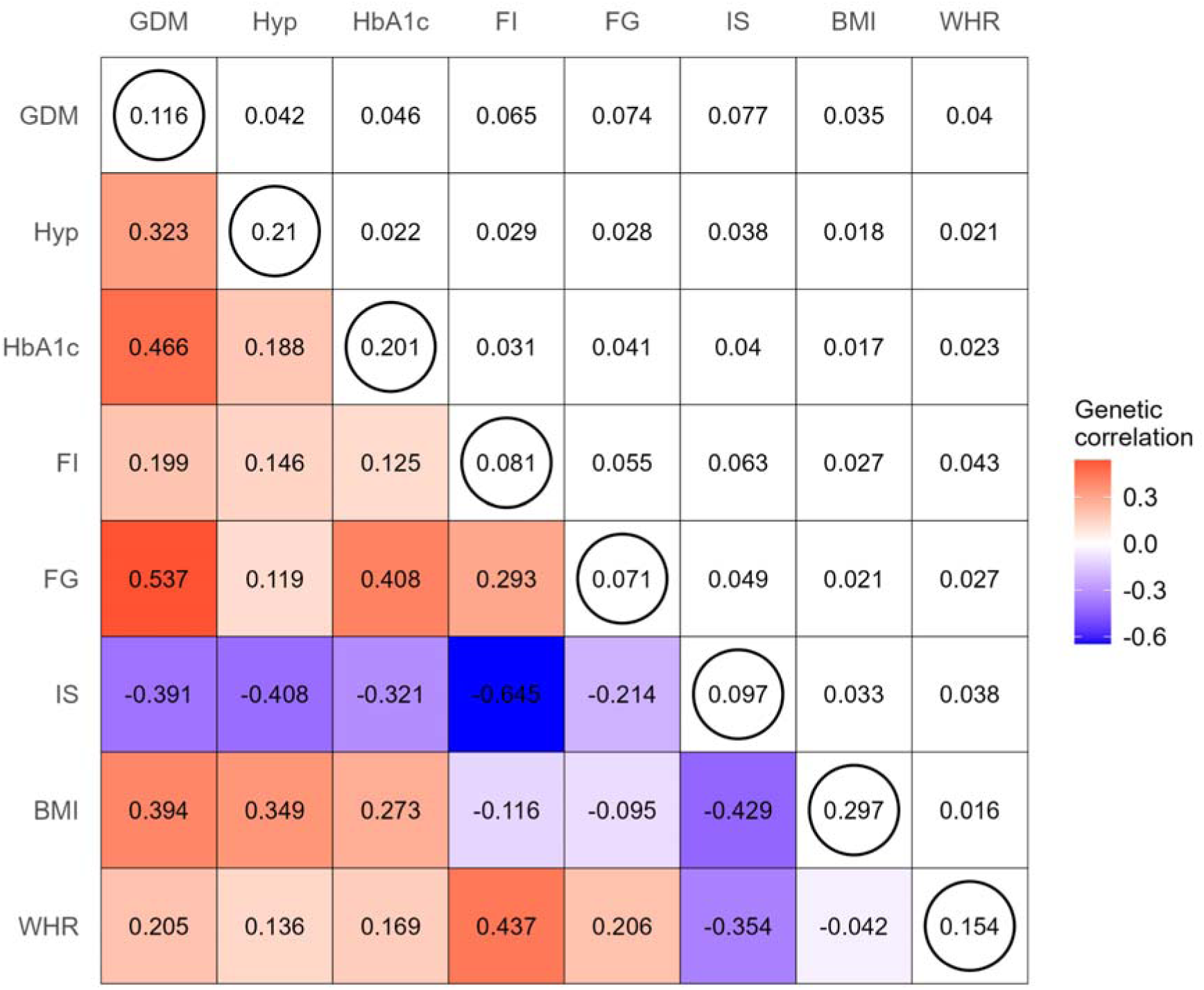
SNP-based heritability and pairwise genetic correlations between the T2D risk factors were estimated using LD score regression. The lower diagonal represents pairwise genetic correlation, the upper diagonal corresponds to standard errors of the genetic correlations and the diagonal circled elements show SNP-based heritability. GDM - gestational diabetes mellitus, Hyp - hypertension, HbA1c - glycated hemoglobin, FI - fasting insulin, FG - fasting glucose, IS - insulin sensitivity, BMI - body mass index, WHR - waist to hip ratio.

### Factor analysis

After estimating the genetic correlations among all pairs of T2D risk factors, EFA revealed a three-factor solution as optimal, accounting for 60.9% of the total variance across eight phenotypes. Based on the EFA findings, we conducted a CFA and the resulting model demonstrated a very good fit with CFI = 0.988 and SRMR = 0.032. The latent factors were clustered and labeled as follows (Figure 2):

1. Glycemic regulation factor - comprising GDM, FG and HbA1c;
2. Insulin resistance and cardiometabolic factor - including FI, IS, Hyp and WHR (adjusted for BMI);
3. Obesity and lifestyle-related factor - encompassing BMI, Hyp, IS, GDM and HbA1c.

**Figure 2.**
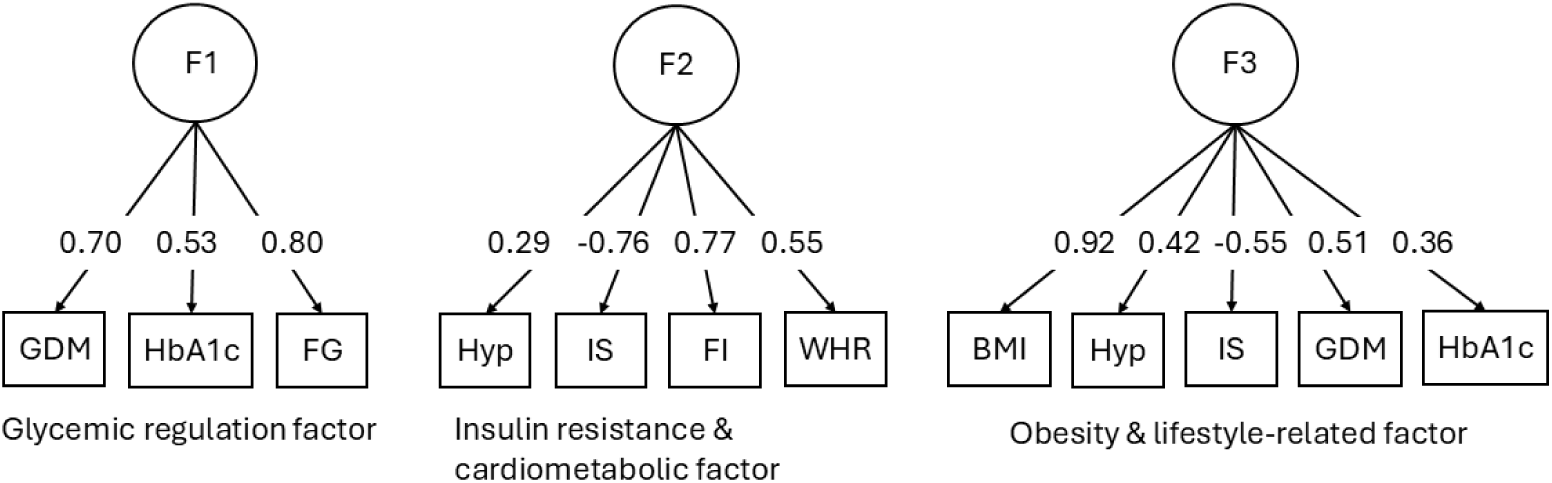
Confirmatory factor analysis results with standardized factor loadings from the Genomic SEM model. F1 - factor 1, F2 - factor 2, F3 - factor 3, GDM - gestational diabetes mellitus, Hyp - hypertension, HbA1c - glycated hemoglobin, FI - fasting insulin, FG - fasting glucose, IS - insulin sensitivity, BMI - body mass index, WHR - waist to hip ratio.

### Multivariate GWAS with Genomic SEM

We performed a multivariate GWAS of the latent factors identified through CFA. This analysis identified numerous genome-wide significant loci, including several well-established signals: GCK in the glycemic regulation factor GWAS (Figure 3a), a gene central to glucose homeostasis; LYPLAL1-AS1 in the insulin resistance and cardiometabolic factor GWAS (Figure 3b), previously associated with insulin-related traits; and FTO in the obesity and lifestyle-related factor GWAS (Figure 3c), a locus known to be linked to obesity.

**Figure 3.**
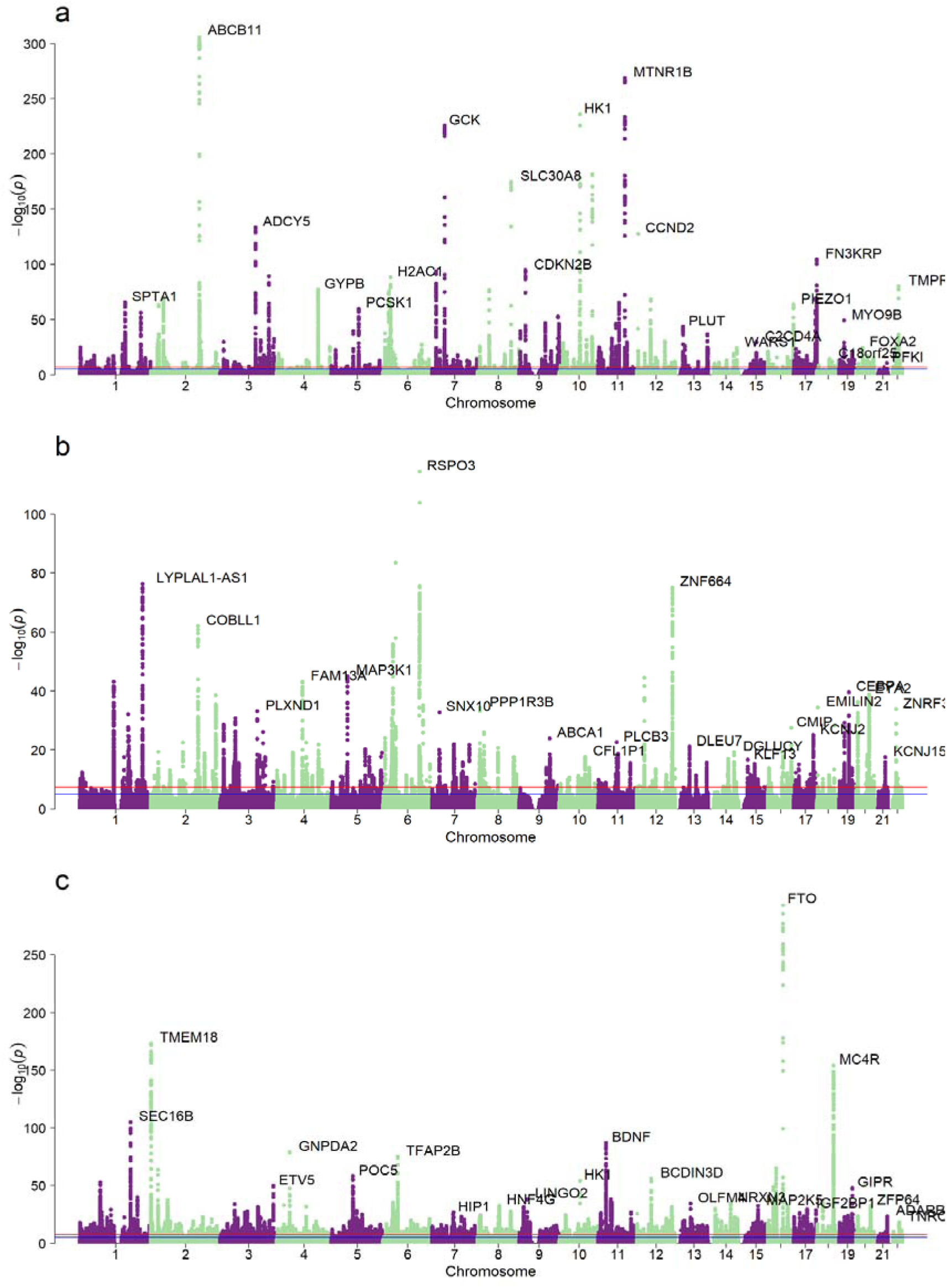
Manhattan plots of multivariate GWAS results. Panel a displays GWAS results for the factor including gestational diabetes, glycated hemoglobin, and fasting glucose. Panel b presents GWAS results for the factor comprising hypertension, insulin sensitivity, fasting insulin, and waist-to-hip ratio. Panel c depicts GWAS results for the factor encompassing body mass index, hypertension, insulin sensitivity, gestational diabetes, and glycated hemoglobin.

### PGS analysis

We computed the genome-wide PGS for latent factors based on multivariate GWAS results. Additionally, PGSs were computed for T2D, FG, FI and BMI to assess how latent factor PGSs relate to T2D genetic risk and to compare them with metabolic PGSs representing the primary phenotypes underlying each factor. We first examined the correlations among the different PGSs. The correlations ranged from 0.01 between the glycemic regulation factor PGS and the insulin resistance & cardiometabolic PGS to 0.95 between the obesity and lifestyle-related PGS and the BMI PGS (Figure 4).

**Figure 4.**
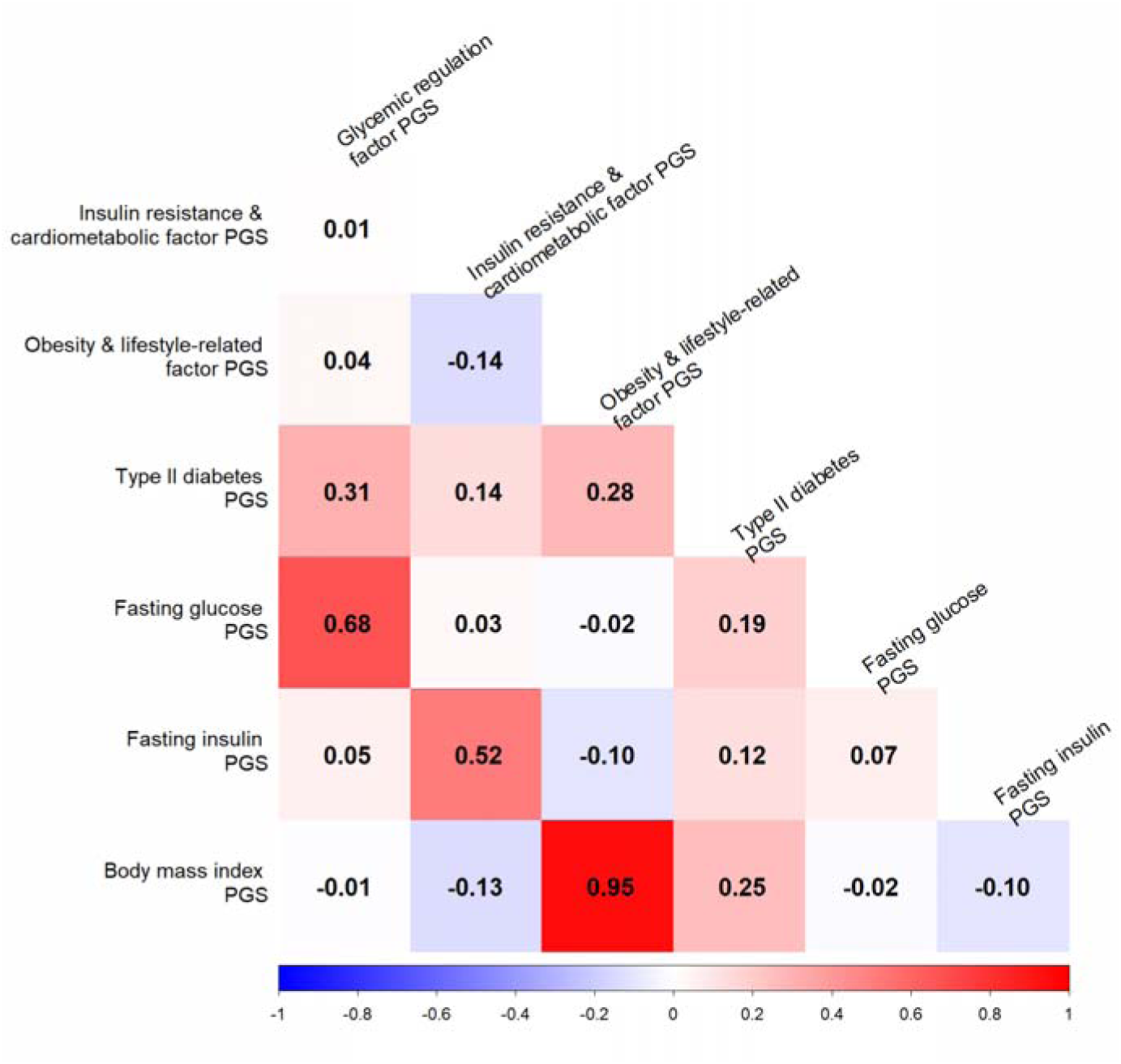
Coefficients of correlation (Pearson) between different polygenic risk scores (PGSs).

Next, we assessed the hazard ratios (HRs) per SD of each PGS for incident T2D in the EstBB cohort. For this we only used incident cases (n=5553) and controls to account for both left-truncation and right-censoring. All PGSs showed significant results, T2D PGSs having expectedly the highest HR per SD (1.45, 95% CI 1.41-1.49) and fasting insulin PGS showing the lowest HR per SD (1.09, 95% CI 1.06-1.12) (Table 1).

**Table 1.** The associations of latent factor PGSs, type II diabetes PGS and fasting glucose, fasting insulin and body mass index PGSs on incident type II diabetes in Cox PH model, adjusted for sex and 10 genetic principal components. The followup starts at the age of joining the biobank and ends at the age of T2D diagnosis.

| PGS | HRs per SD (95% CI) | Pvalue |
| --- | --- | --- |
| Glycemic regulation factor PGS | 1.29 (1.25; 1.33) | $6.28 \cdot 10^{-76}$ |
| Insulin resistance and cardiometabolic factor PGS | 1.16 (1.12; 1.20) | $1.67 \cdot 10^{-25}$ |
| Obesity & lifestyle-related factor PGS | 1.31 (1.27; 1.35) | $7.39 \cdot 10^{-89}$ |
| Type II diabetes PGS | 1.45 (1.41; 1.49) | $9.27 \cdot 10^{-164}$ |
| Fasting glucose PGS | 1.18 (1.15; 1.22) | $1.60 \cdot 10^{-35}$ |
| Fasting insulin PGS | 1.09 (1.06; 1.12) | $3.72 \cdot 10^{-10}$ |
| Body mass index PGS | 1.31 (1.27; 1.35) | $7.79 \cdot 10^{-88}$ |

Furthermore, we evaluated the overlap between the T2D PGS and the latent factor PGSs. Among EstBB participants with high T2D PGS, nearly 70% also had at least one elevated latent factor PGS (Figure 5).

**Figure 5.**
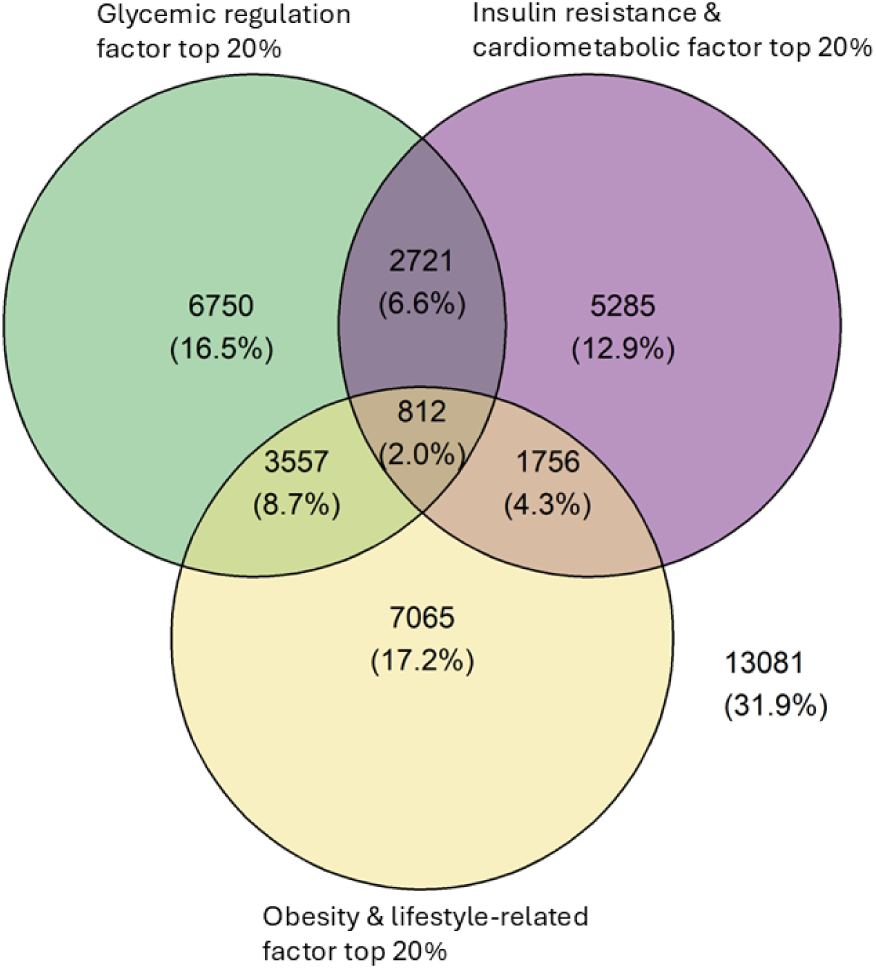
Distribution of high latent factor PGSs among people in the Estonian Biobank, who have also high PGS for type II diabetes.

### PheWAS with PGS

To investigate the broader health impacts associated with the latent factors, we conducted a PGS–PheWAS in 118,628 EstBB participants. Among the latent factor polygenic scores (PGSs), the obesity and lifestyle-related factor PGS exhibited associations with a broader range of health outcomes compared with the glycemic regulation and insulin resistance and cardiometabolic factor PGSs.

The glycemic regulation factor PGS showed strong associations with T2D (ICD-10 code E11), elevated blood glucose levels (R73), and gestational diabetes (O24) (Figure 6a). The insulin resistance and cardiometabolic factor PGS demonstrated its strongest associations with T2D, hypertension (I10, I11), and disorders of lipoprotein metabolism (E78) (Figure 6b). In contrast, the obesity and lifestyle-related factor PGS was most strongly associated with obesity (E66), T2D, and hypertension (Figure 6c).

**Figure 6.**
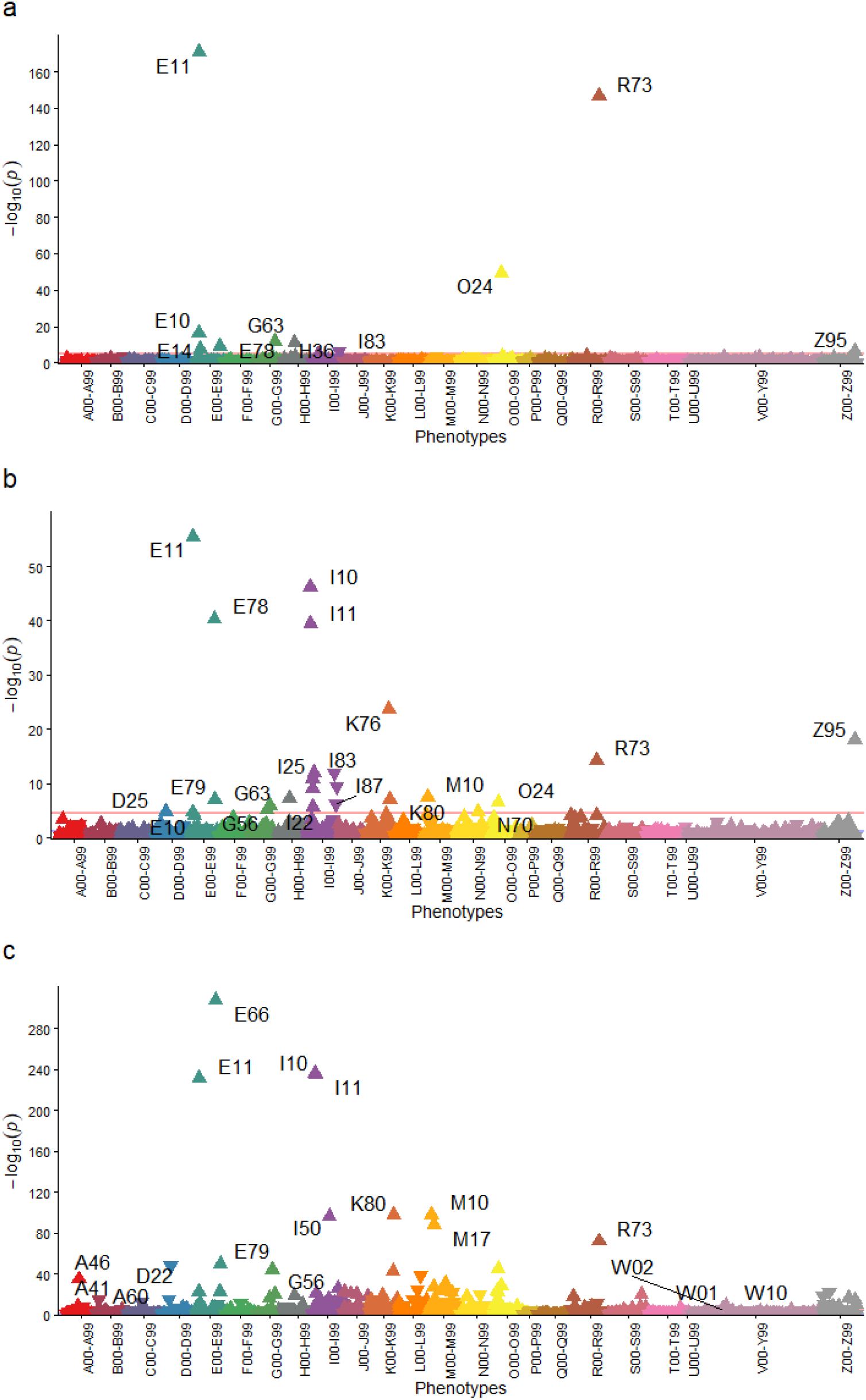
Phewas results for latent factor PGSs. Panel **a** corresponds to glycemic regulation factor PGS, panel **b** to insulin resistance and cardiometabolic factor PGS and panel **c** to obesity and lifestyle-related factor PGS results.

For comparison, we also performed PGS–PheWAS analyses using PGSs for fasting glucose (FG), fasting insulin (FI), and body mass index (BMI). These results largely mirrored those observed for the latent factor PGSs, particularly the close correspondence between the obesity and lifestyle-related factor PGS and the BMI PGS. Notably, however, the latent factor PGSs captured a broader spectrum of comorbidities overall (Figure 7a–c). The top associations for FG, FI, and BMI PGSs were generally similar to those observed for their corresponding latent factors, with the exception of FG, which showed its strongest association with elevated blood glucose levels, whereas the glycemic regulation factor PGS was most strongly associated with T2D.

**Figure 7.**
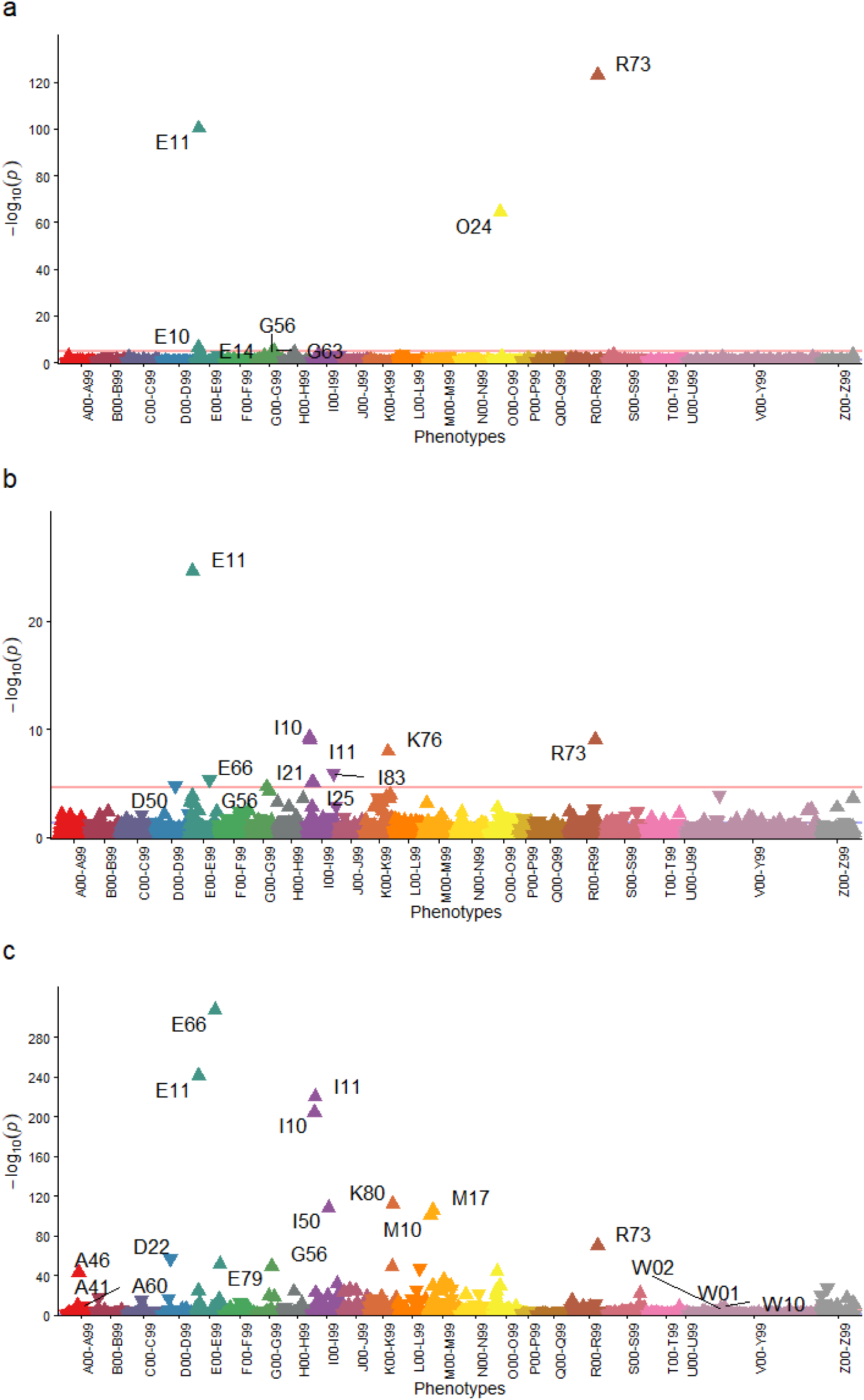
Phewas results for fasting glucose, fasting insulin and body mass index PGS. Panel **a** corresponds to fasting glucose PGS, panel **b** to fasting insulin PGS and panel **c** to body mass index PGS results.

### Comorbidity analyses

Each latent factor PGS was associated with a specific subset of T2D-related comorbidities, with some showing a protective effect for certain conditions (Figure 8). The number of events for comorbidities ranged from 59 for glomerular disorders in diseases classified elsewhere to 3570 for disorders of lipoprotein metabolism and other lipidaemias (for details see Supplementary Table 2). Among the three latent factors, the obesity and lifestyle-related factor PGS was associated with the broadest spectrum of comorbidities.

**Figure 8.**
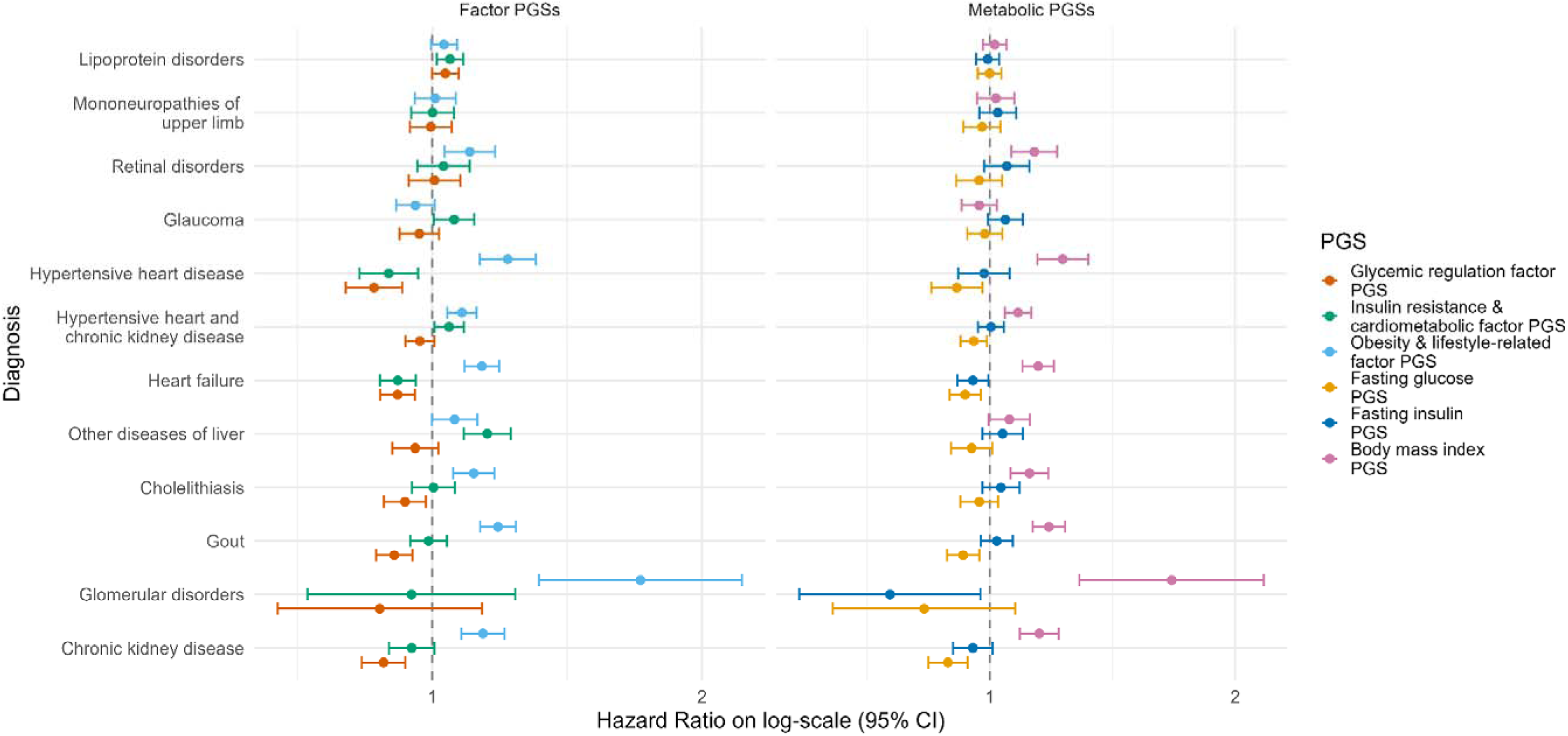
Forest plot of hazard ratios (HRs) per SD with 95% CI from Cox PH model for incidence of comorbidites after type 2 diabetes (T2D) diagnosis. Cox PH followup starts at the age of T2D diagnosis, ends at the age of comorbidity diagnosis, and the model is adjusted for sex and 10 genetic principal components.

We compared the results to metabolic PGSs (FG, FI and BMI PGS) and found that latent factor PGSs generally showed stronger associations. The C-statistics for factor PGSs ranged from 0.540 to 0.751, while those for metabolic PGSs ranged from 0.540 to 0.745 (Supplementary Table 3). Notably, for several comorbidities metabolic PGSs showed no significant association, while the corresponding latent factor PGSs demonstrated clear relationship with the comorbidity.

The survival curves stratified by PGS category (high, medium, low) further illustrated that these PGSs effectively distinguish differences in comorbidity incidence over time (Figures 9–11).

**Figure 9.**
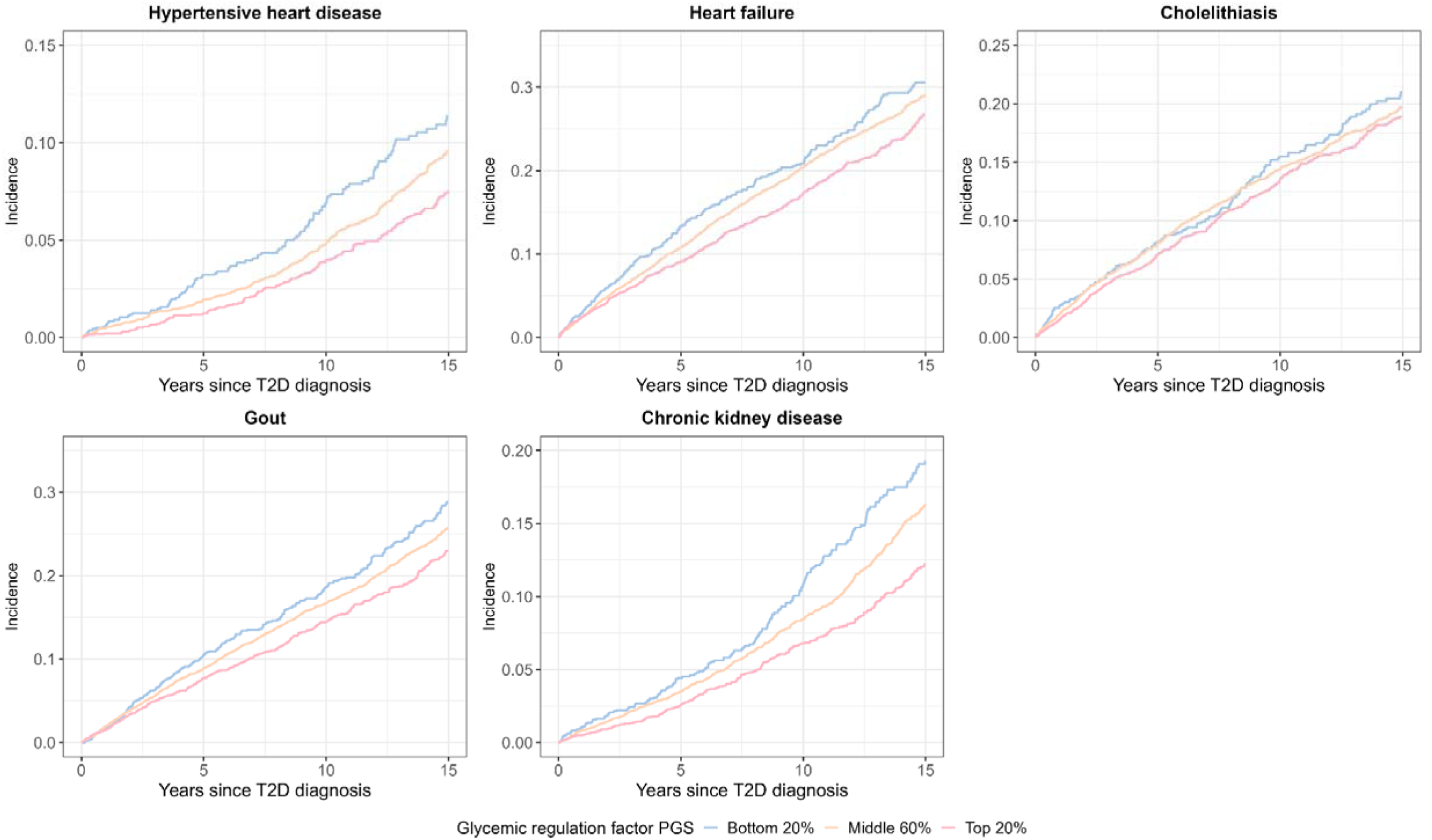
Kaplan-Meier curves for the incidence of T2D comorbidities that showed significant association with glycemic regulation factor PGS in Cox PH model.

**Figure 10.**
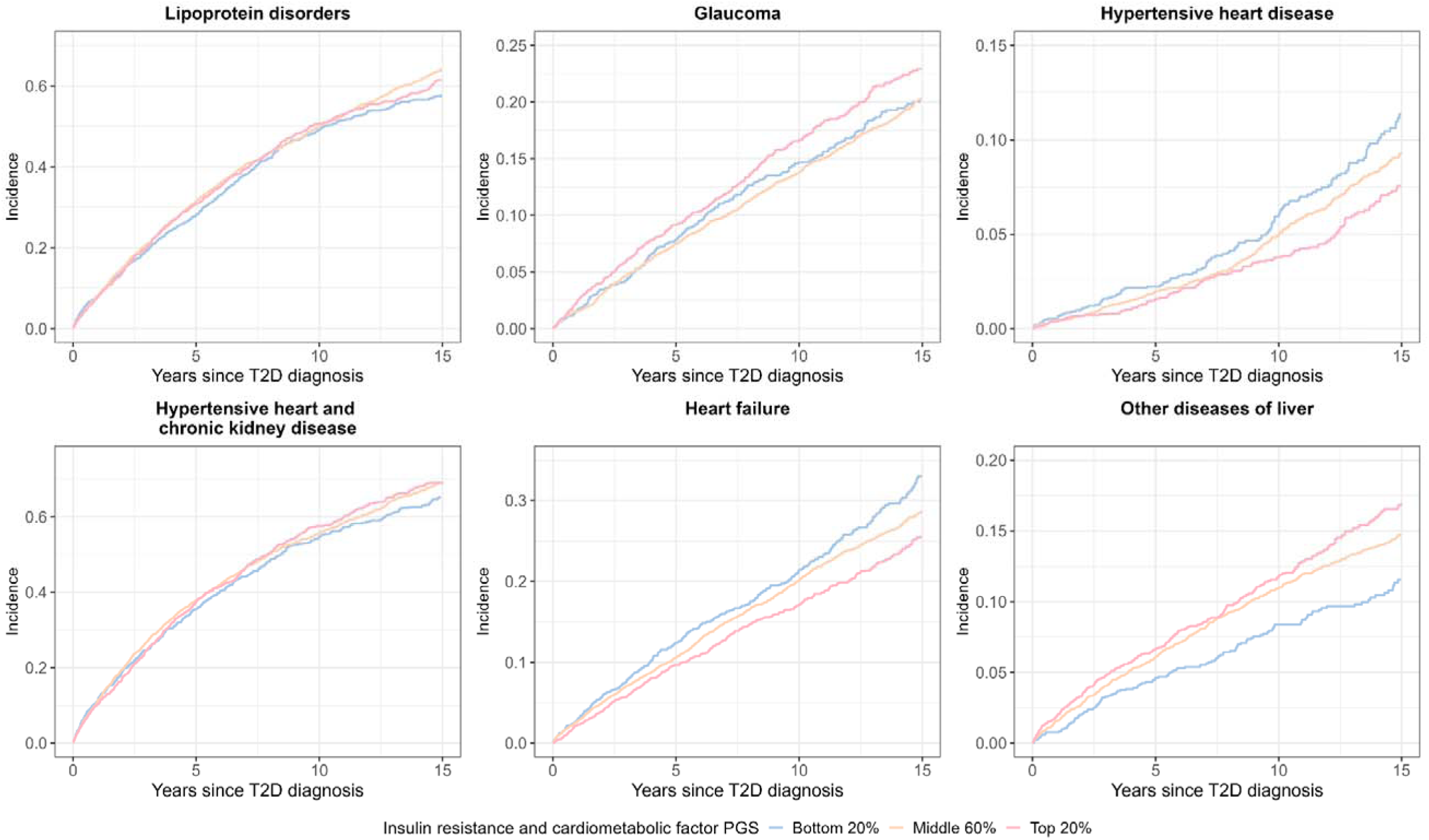
Kaplan-Meier curves for the incidence of T2D comorbidities that showed significant association with insulin resistance and cardiometabolic factor PGS in Cox PH model.

**Figure 11.**
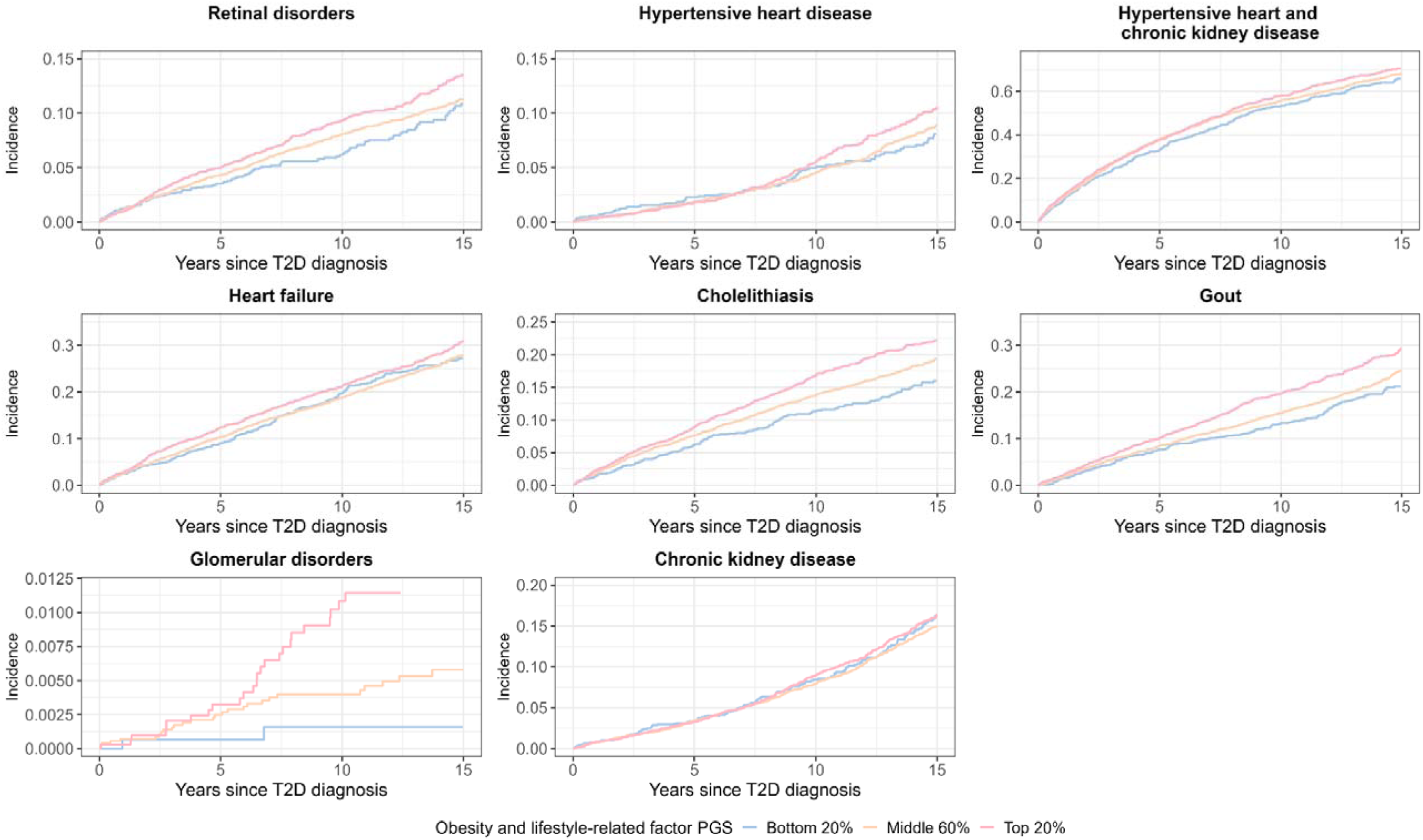
Kaplan-Meier curves for the incidence of T2D comorbidities that showed significant association with obesity and lifestyle-related risk factor PGS in Cox PH model

## Discussion

In this study, we used genomic SEM to characterise the shared genetic architecture of T2D and major upstream cardiometabolic traits. We identified three latent genetic factors representing glycaemic regulation, insulin resistance with cardiometabolic risk, and obesity and lifestyle-related liability. PGSs derived from multivariate GWAS of these latent factors were associated with incident T2D and revealed distinct, factor-specific patterns of comorbidity. Notably, the obesity and lifestyle-related factor showed the broadest phenotypic impact across both population-wide PheWAS and post-T2D comorbidity analyses. Furthermore, a large proportion of individuals with high overall T2D polygenic risk also exhibited elevation in one or more latent factor polygenic scores, enabling decomposition of aggregate genetic risk into biologically interpretable components.

The glycaemic regulation factor, defined by positive loadings on fasting glucose, HbA1c, and gestational diabetes, reflects shared genetic liability related to glucose homeostasis and β-cell function. This factor corresponds closely to β-cell–dysfunction–dominant clusters identified in prior genetic and phenotypic classification approaches and aligns with genetic pathways previously implicated in insulin secretion defects and early dysglycaemia, which are central to T2D development (5,6). Its association with incident T2D supports the long-standing view that impaired glycaemic control constitutes a primary disease axis. However, compared with other factors, this latent dimension showed a narrower comorbidity profile, suggesting that while glycaemic dysregulation is critical for disease onset, it may play a more limited role in shaping broader systemic complications once T2D is established.

The insulin resistance and cardiometabolic factor is defined by positive loadings on fasting insulin, hypertension, and body fat distribution, and a negative loading on insulin sensitivity, consistent with increased insulin resistance. It closely parallels insulin-resistant diabetes subtypes as well as lipodystrophy- and liver/lipid–related clusters identified in prior genetic and clinical clustering studies, which exhibit elevated risks of cardiovascular and hepatic disease (9,40). The comorbidity patterns observed here, including associations with liver and cardiovascular outcomes, are consistent with longitudinal studies showing increased burden of these conditions in insulin-resistant diabetes groups (41). These findings reinforce the interpretation that insulin resistance represents a genetically distinct pathway that contributes not only to T2D risk but also to downstream cardiometabolic morbidity.

The obesity and lifestyle-related factor was characterised by positive loadings on body mass index, hypertension, and glycaemic traits, and a negative loading on insulin sensitivity, consistent with a broad metabolic liability extending beyond adiposity. This factor demonstrated the widest range of associations across PGS-PheWAS and comorbidity analyses, consistent with obesity cluster in prior genetic clustering studies showing that this cluster affects the largest number of T2D comorbidities and often with the strongest effect sizes (5,6,42). The breadth of associations observed here suggests that genetic susceptibility to obesity-related metabolic stress exerts pervasive effects across multiple organ systems, likely through chronic inflammation, ectopic fat accumulation, and long-term cardiometabolic burden. The strong concordance between our findings and previous partitioned PGS analyses further supports the robustness of this pathway.

These findings have several important implications. First, they demonstrate that T2D genetic risk is not monolithic but instead reflects multiple biologically distinct pathways that can be captured using multivariate genetic modelling. Latent factor PGS provide mechanistic resolution beyond conventional disease-level PGS, enabling stratification of individuals according to dominant underlying drivers of risk. This has potential relevance for precision prevention, targeted surveillance, and early intervention strategies.

Second, the strong and pervasive impact of the obesity and lifestyle-related factor highlights the importance of addressing adiposity-driven pathways in both population-level prevention and post-diagnosis management. The observation that many individuals with high T2D genetic risk carry elevated risk across multiple latent factors suggests that combined or pathway-informed risk profiling may be more informative than single-score approaches.

Future work should extend these analyses to non-European populations, integrate environmental and lifestyle exposures, and evaluate whether latent factor polygenic scores improve clinical prediction beyond established risk models. Longitudinal studies examining treatment response and progression across genetically defined subgroups may further clarify how these latent pathways translate into actionable clinical stratification.

A key strength of this study is the use of genomic structural equation modelling to jointly analyse correlated cardiometabolic traits, allowing latent biological dimensions of T2D risk to emerge directly from genetic data. The integration of multivariate GWAS with longitudinal biobank data enabled both genetic discovery and clinically meaningful downstream analyses.

Genomic SEM requires all input GWASs to be derived from the same ancestry because differences in LD structure across populations can bias the estimation of genetic covariance; consequently, our analyses were restricted to European-ancestry populations, limiting generalisability. In addition, we were unable to use GWASs that included EstBB participants in most cases to avoid sample overlap; however, in one instance we selected a GWAS that included EstBB and subsequently excluded approximately 10,000 overlapping EstBB participants who had contributed to fasting glucose and fasting insulin GWASs from downstream analyses. This exclusion likely reduced statistical power and may have attenuated some associations. Finally, while latent factor polygenic scores capture genetic liability, they do not account for environmental or behavioural factors that substantially modify T2D risk and progression.

In conclusion, this study shows that the genetic architecture of T2D can be decomposed into three latent factors reflecting glycaemic regulation, insulin resistance with cardiometabolic risk, and obesity and lifestyle-related liability. These factors exhibit distinct associations with T2D onset and comorbidity profiles, with obesity-related genetic liability showing the broadest systemic impact. By linking aggregate T2D genetic risk to specific biological pathways, this work provides a framework for more nuanced genetic risk stratification and supports the use of multivariate approaches to advance precision medicine in T2D.

## Data Availability

All data produced in the present study are available upon reasonable request to the authors

## Acknowledgements

MK, KL, MM and RM have been supported by the Estonian Research Council grant PRG1911. MK and KF have been supported by the Estonian Research Council grants PRG1197 and PRG3105. MK, KF and RM were supported by the Ministry of Education and Research Centres of Excellence grant TK214. KL, RM and KF have received funding from the European Union’s Horizon Europe research and innovation programme under grant agreement No 101060011. Views and opinions expressed are however those of the author(s) only and do not necessarily reflect those of the European Union or European Research Executive Agency. Neither the European Union nor the granting authority can be held responsible for them. APM acknowledges support from the NIHR Manchester Biomedical Research Centre (NIHR203308).

## Ethics

The activities of the EstBB are regulated by the Human Genes Research Act, which was adopted in 2000 specifically for the operations of the EstBB. Individual level data analysis in the EstBB was carried out under ethical approval 1.1-12/624 from the Estonian Committee on Bioethics and Human Research (Estonian Ministry of Social Affairs), using data according to release application 6-7/GI/1566 from the Estonian Biobank.

